# The economic burden of RSV-associated illness in children aged <5 years, South Africa 2011-2016

**DOI:** 10.1101/2022.06.20.22276632

**Authors:** Jocelyn Moyes, Stefano Tempia, Sibongile Walaza, Meredith L. McMorrow, Florette Treurnicht, Nicole Wolter, Anne von Gottberg, Kathleen Kahn, Adam L Cohen, Halima Dawood, Ebrahim Variava, Cheryl Cohen

## Abstract

**Introduction:** Data on the economic burden of RSV-associated illness will inform decisions on the programmatic implementation of maternal vaccines and monoclonal antibodies. We estimated these costs in fine age bands to allow more accurate cost-effectiveness models to account for limited duration of protection conferred by short or long acting interventions.

**Methods:** We conducted a costing study at sentinel sites across South Africa to estimate out-of-pocket and indirect costs for RSV-associated mild and severe illness. We collected facility-specific costs for staffing, equipment, services, diagnostic tests and treatment. Using case-based data we calculated a patient day equivalent (PDE) for RSV-associated hospitalisations or clinic visits; the PDE was multiplied by the number of days of care to provide a case-cost to the healthcare system. We estimated the costs in 3-month age intervals in children aged <1 years and as a single group for children aged 1-4 years. We then applied our data to a modified version of the World Health Organization tool for estimating mean annual national cost burden, including medically and non-medically attended RSV-associated illness.

**Results:** The estimated mean annual cost of RSV-associated Illness in children aged <5 years was United States dollars ($)137 204 393, of which 81% ($111 742 713) were healthcare system incurred, 6% ($8 881 612) were out of pocket expenses and 13% ($28 225 801) were indirect costs. Thirty-three percent ($45 652 677/$137 204 393) of the total cost in children aged <5 years was in the <3-month age group, of which 52% ($71 654 002) were healthcare system incurred. The costs of non-medically attended cases increased with age from $3 307 218 in the <3-month age group to $8 603 377 in the 9-11-month age group.

**Conclusion:** Among children <5 years of age with RSV in South Africa, the highest cost burden was in young infants; therefore, interventions against RSV targeting this age group are important to reduce the health and cost burden of RSV-associated illness.

## Introduction

Globally, an estimated 33 million RSV-associated lower respiratory tract infections (LRTI) and 118 000 deaths occur annually in children under 5 years of age^1^. The only currently licensed Respiratory Syncytial Virus (RSV)-prevention method is a short acting monoclonal antibody (MAB) (palivizumab) injected monthly, usually recommend for high-risk infants due to cost. MAB is not widely available in many low- and middle-income settings such as South Africa. However, a number of new technologies, including maternal vaccines and longer acting monoclonal antibodies, are being developed to prevent RSV-associated illness in children^2^.

Global cost effectiveness models based on estimates of burden in low- and middle-income countries suggest up to 1,2 million cases, 104 million hospital admissions and 3,000 deaths could be averted annually through maternal RSV vaccination, including saving 186 million United States Dollars ($) in healthcare costs^3^. MAB strategies could prevent more even cases, with more cost savings than a maternal vaccine ^3^. In a recent global analysis, cost effectiveness was modeled in two age bands, <1 year and <5 years. These broad age bands do not account for the fact that MAB and maternal vaccines in trials may have a limited duration of effect (up to six months) within the <1-year age band. Cost effectiveness models may be improved by considering finer age bands in infants, specifically in infants <6 months of age. Li et al, for their model of cost effectiveness, list the following as important parameters to quantify for country-level cost-effectiveness models; cost of hospitalized and non-medically attended cases and mortality burden, to ensure cost effectiveness models are accurate and specific.^3^

In South Africa the burden of RSV-associated mild and severe illness is substantial. From 2012-2016, the mean annual number of RSV-associated severe cases in children aged <5 years was 96 220 (95% confidence interval (CI) 66 470-132 844) with 78 571 (95% CI 56 187-105 831) severe cases in children aged <1 year. In the same period the mean number of RSV-associated deaths in children aged <5 years was 650 (95% CI 475-947) (ref burden paper, in clearance). There were also a large number of non-medically attended cases for both severe and mild RSV-associated illness (204080 95% CI 103191-341183 and 51605 95% CI 33739-75306) respectively (ref burden paper in clearance). With these detailed burden estimates, we can describe cost burden of RSV-associated illness to inform cost effectiveness models.

We estimate the costs and cost burden associated with RSV-associated mild and severe illness in in children aged <5 years (in 3-month age groups from 0-11 months and then in those aged 1-4 years in South Africa from 2011 through 2016. Specifically, healthcare system costs, out of pocket expenses to the caregivers of children, the indirect cost and cost of non-medically attended illness.

## Methods

We collected all costs in South Africa Rands (ZAR) and converted to United States Dollars ($) using the average monthly ZAR to $ exchange in 2014 based on the South African all items consumer price index^4^.

We conducted a costing study at sentinel surveillance sites (5 hospitals and 2 public health clinics in 4 of South Africa’s 9 provinces) that are part of a national surveillance programme for pneumonia to estimate out-of-pocket and indirect costs incurred by caregivers of children with RSV-associated mild (influenza-like illness) and severe illness (severe acute respiratory illness (SARI)) from January through December 2014 providing a baseline of costing estimates (and adjusted for consumer price index for other years). We enrolled 2 severe cases (hospitalised) and 2 mild (outpatient influenza-like illness) cases per week in each of the following age groups: <1 year and 1-4 years. Data were collected by structured interview including caregiver out-of-pocket costs for the visit or admission. These included the cost of transport, food, over-the-counter medication used and cost of any private practitioner or traditional healer visits prior to study clinic visits or hospitalisation. Indirect costs including loss of income were also collected from the caregiver. We applied the national minimum wage estimates to the days of lost income for the caregiver and to days of hospitalization. Minimum wage during the study period was $1.3 per day ^5,6^.

Using case-based data we calculated a patient day equivalent (PDE) for RSV-associated hospitalisations, which was multiplied by the number of days of care to provide a case-cost to the healthcare system. All hospitalization costs were assigned to the healthcare facility as healthcare is free of charge to children aged <5 years in South Africa. Itemized hospital costs and procedures (cost of facility services, administration, laundry, furniture, physician and nurse consultations, intensive care fees, chest X-rays, laboratory tests and oxygen therapy) were obtained from the National Department of Health (NDoH) and the state price list of the National Health Laboratory Service (NHLS)^7,8^. A similar exercise estimated outpatient related healthcare system costs.

Direct medical costs outside of public facilities including over-the-counter (OTC) medications, consultation with traditional healers and private practitioners, direct non-medical costs (like transportation) and indirect costs (loss of income) were obtained from a community survey conducted in 2013 in 4 of 9 provinces (baselines estimates) and adjusted for consumer price index ^4^.

To estimate the national economic burden of RSV-associated illness, 2013 to 2015 data were entered into a modified version of the World Health Organization tool for estimating the mean annual national cost burden, including medically and non-medically attended RSV-associated illness^9^ (burden paper). We modified the model to include the direct and indirect cost estimates for caregivers for non-medically attended mild and severe illness as approximately 70% of mild and 50% of severe RSV-associated cases in children aged <5 years are not medically attended in our setting (ref burden paper, recently cleared by CDC and due for submission to a journal).

Population data were obtained from Statistics South Africa as a mean annual population for 2011-2016 and case fatality ratios were obtained from surveillance data during the same time period^10^. We estimated the cost and cost burden of RSV-associated illness in 3-month age intervals in children aged <1 years and as a single group for children aged 1-4 years.

We calculated years of life lost by applying the life expectancy estimates for South Africa in 2016 to the years lost for deaths in each age group^11^.

## Results

### Costing study

We enrolled 527 children with mild illness and 675 children with severe illness into the costing study. The detection rate of RSV was 10% (57) in those with mild illness and 26% (178) in those with severe illness. The mean annual household income including government grants was $559 ($ 46.58 monthly) and the median was $475 (2.5%-97.5% range $191-$1156)^12^.

### Healthcare system costs

Healthcare system costs were highest for the youngest infants (aged 0-2 months) at $921.66 per episode as compared with $623.59 for infants aged 3-5 months, $583.71 for infants aged 6-8 months, $716.68 for infants aged 9-11 months and $591.44 for children aged 1-4 years. This cost is driven by a mean duration of hospitalization 6.2 days in infants aged 0-2 months as compared to 4.7 days, 4.4 days, 5.6 days and 4.1 days in children aged 3-5 months, 6-8 months, 9-11 months, or 12-59 months, respectively. Healthcare system costs for mild illness were more uniform; ranging from $24.98 among infants aged 0-2 months to $25.66 in children aged 12-59 months per episode (Table 1).

**Table 1:** The mean cost per episode of Respiratory Syncytial Virus (RSV)-associated mild and severe illness in children aged <5 years, South Africa, 2014

| RSV-associated Severe Illness |  |  |  |  |  |  |  |  |  |  |  |  |  |  |  |  |
| --- | --- | --- | --- | --- | --- | --- | --- | --- | --- | --- | --- | --- | --- | --- | --- | --- |
| Age group in months |  | 0-2 |  |  | 3-5 |  |  | 6-8 |  |  | 9-11 |  |  | 12-59 |  |  |
| Item | Unit cost (\$) | Proportion of children tested or requiring treatment | Duration of incurred cost (days) | Mean cost per episode (\$) | Proportion of children requiring treatment | Duration of incurred cost (days) | Mean cost per episode | Proportion of children requiring treatment | Duration of incurred cost (days) | Mean cost per episode (\$) | Proportion of children requiring treatment | Duration of incurred cost (days) | Mean cost per episode (\$) | Proportion of children requiring treatment | Duration of incurred cost (days) | Mean cost per episode (\$) |
| Facility fee (24H) | 84,20 | 1,00 | 6,24 | 525,11 | 1,00 | 4,74 | 399,37 | 1,00 | 4,37 | 367,66 | 1,00 | 5,60 | 471,50 | 1,00 | 4,13 | 347,61 |
| Consultation (24H) | 24,29 | 1,00 | 6,24 | 151,46 | 1,00 | 4,74 | 115,20 | 1,00 | 4,37 | 106,05 | 1,00 | 5,60 | 136,00 | 1,00 | 4,13 | 100,27 |
| ICU (24H) | 691,61 | 0,03 | 7,00 | 165,41 | 0,01 | 3,83 | 25,80 | 0,02 | 0,67 | 10,97 | 0,02 | 0,67 | 8,55 | 0,04 | 2,00 | 51,48 |
| Chest X-ray | 26,16 | 0,58 | 1,00 | 15,09 | 0,70 | 1,00 | 18,22 | 0,85 | 1,00 | 22,36 | 0,83 | 1,00 | 21,69 | 0,74 | 0,86 | 16,58 |
| Oxygen (24H) | 34,29 | 0,45 | 1,20 | 18,44 | 0,44 | 1,20 | 18,23 | 0,44 | 1,20 | 18,23 | 0,53 | 1,20 | 21,84 | 0,91 | 1,03 | 32,06 |
| Antibiotic treatment | 18,30 | 0,96 | 1,00 | 17,64 | 0,93 | 1,00 | 17,06 | 0,93 | 1,00 | 16,95 | 0,92 | 1,00 | 16,83 | 0,96 | 0,86 | 15,04 |
| Other medications | 8,04 | 1,00 | 1,00 | 8,04 | 1,00 | 1,00 | 8,04 | 1,00 | 1,00 | 8,04 | 1,00 | 1,00 | 8,04 | 1,00 | 0,86 | 6,89 |
| HIV rapid | 5,36 | 0,00 | 1,00 | 0,00 | 0,29 | 1,00 | 1,54 | 0,58 | 1,00 | 3,09 | 0,00 | 1,00 | 0,00 | 0,00 | 0,86 | 0,00 |
| HIV ELISA | 4,38 | 0,15 | 1,00 | 0,64 | 0,25 | 1,00 | 1,08 | 0,04 | 1,00 | 0,19 | 0,07 | 1,00 | 0,32 | 0,24 | 0,86 | 0,90 |
| HIV PCR | 30,54 | 0,28 | 1,00 | 8,43 | 0,23 | 1,00 | 7,02 | 0,44 | 1,00 | 13,58 | 0,48 | 1,00 | 14,76 | 0,31 | 0,86 | 8,03 |
| HIV viral load | 27,23 | 0,00 | 1,00 | 0,11 | 0,01 | 1,00 | 0,27 | 0,00 | 1,00 | 0,00 | 0,02 | 1,00 | 0,50 | 0,05 | 0,86 | 1,28 |
| HIV CD4+ | 14,55 | 0,00 | 1,00 | 0,03 | 0,01 | 1,00 | 0,09 | 0,00 | 1,00 | 0,00 | 0,00 | 1,00 | 0,00 | 0,04 | 0,86 | 0,47 |
| Bacterial culture | 4,11 | 0,89 | 1,00 | 3,65 | 0,68 | 1,00 | 2,80 | 0,83 | 1,00 | 3,40 | 0,69 | 1,00 | 2,85 | 0,59 | 0,86 | 2,09 |
| Tuberculosis test | 25,00 | 0,01 | 1,00 | 0,22 | 0,01 | 1,00 | 0,20 | 0,04 | 1,00 | 1,12 | 0,09 | 1,00 | 2,37 | 0,07 | 0,86 | 1,59 |
| Blood cell count | 4,64 | 0,58 | 1,00 | 2,71 | 0,57 | 1,00 | 2,66 | 1,00 | 1,00 | 4,64 | 0,83 | 1,00 | 3,87 | 0,24 | 0,86 | 0,96 |
| CRP test | 5,89 | 0,47 | 1,00 | 2,77 | 0,70 | 1,00 | 4,11 | 0,85 | 1,00 | 5,01 | 0,80 | 1,00 | 4,69 | 0,83 | 0,86 | 4,20 |

| ESR test | 2,77 | 0,07 | 1,00 | 0,19 | 0,13 | 1,00 | 0,37 | 0,18 | 1,00 | 0,51 | 0,32 | 1,00 | 0,87 | 0,17 | 0,86 | 0,41 |
| --- | --- | --- | --- | --- | --- | --- | --- | --- | --- | --- | --- | --- | --- | --- | --- | --- |
| Urea and creatinine | 2,41 | 0,72 | 1,00 | 1,73 | 0,64 | 1,00 | 1,54 | 0,80 | 1,00 | 1,93 | 0,83 | 1,00 | 1,99 | 0,77 | 0,86 | 1,58 |
| Cost per episode |  |  |  | 921,66 |  |  | 623,59 |  |  | 583,71 |  |  | 716,68 |  |  | 591,44 |
| Patient day equivalent (PDE) |  |  |  | 147,70 |  |  | 131,56 |  |  | 133,57 |  |  | 127,98 |  |  | 143,26 |
| **RSV-associated mild illness** |  |  |  |  |  |  |  |  |  |  |  |  |  |  |  |  |
| | Unit cost | Proportion of children requiring treatment | Duration of incurred cost (days) | Mean cost per episode (\$) | Proportion of children requiring treatment | Duration of incurred cost (days) | Mean cost per episode (\$) | Proportion of children requiring treatment | Duration of incurred cost (days) | Mean cost per episode (\$) | Proportion of children requiring treatment | Duration of incurred cost (days) | Mean cost per episode (\$) | Proportion of children requiring treatment | Duration of incurred cost (days) | Mean cost per episode (\$) |
| Facility fee | 8,4 | 1 | 1 | 8,4 | 1 | 1 | 8,4 | 1 | 1 | 8,4 |  | 1 | 8,4 | 1 | 1 | 8,4 |
| Consultation | 8,9 | 1 | 1 | 8,9 | 1 | 1 | 8,9 | 1 | 1 | 8,9 |  | 1 | 8,9 | 1 | 1 | 8,9 |
| Antibiotic treatment | 11,3 | 0,39 | 1 | 4,41 | 0,39 | 1 | 4,41 | 0,41 | 1 | 4,63 | 0,42 | 1 | 4,75 | 0,45 | 1 | 5,09 |
| Other medication | 3,3 | 0,99 | 1 | 3,27 | 0,99 | 1 | 3,27 | 0,99 | 1 | 3,27 | 0,99 | 1 | 3,27 | 0,99 | 1 | 3,27 |
| Total |  |  |  | 24,98 |  |  | 24,98 |  |  | 25,20 |  | 1 | 25,29 |  |  | 25,66 |

### Cost to caregiver

For children with severe illness the cost to caregivers was highest in the 0-2-month age group (total out of pocket and indirect costs) at $77.55 as compared to $62.21 for infants 3-5 months, $51.27 for infants 6-8 months, $72.19 for infants 9-11 months and $66.77 for children. For children admitted with severe illness, the cost to the caregiver was between 110% ($51.27/$46.58) and 166% ($77.55/$46.58) of the mean monthly household income. Caregiver costs were similar across the age groups for children with mild illness (range $11.28-$11.99) (Table 2) and were an additional 26% of the mean annual household income ($11.99/$46.58).

**Table 2:**
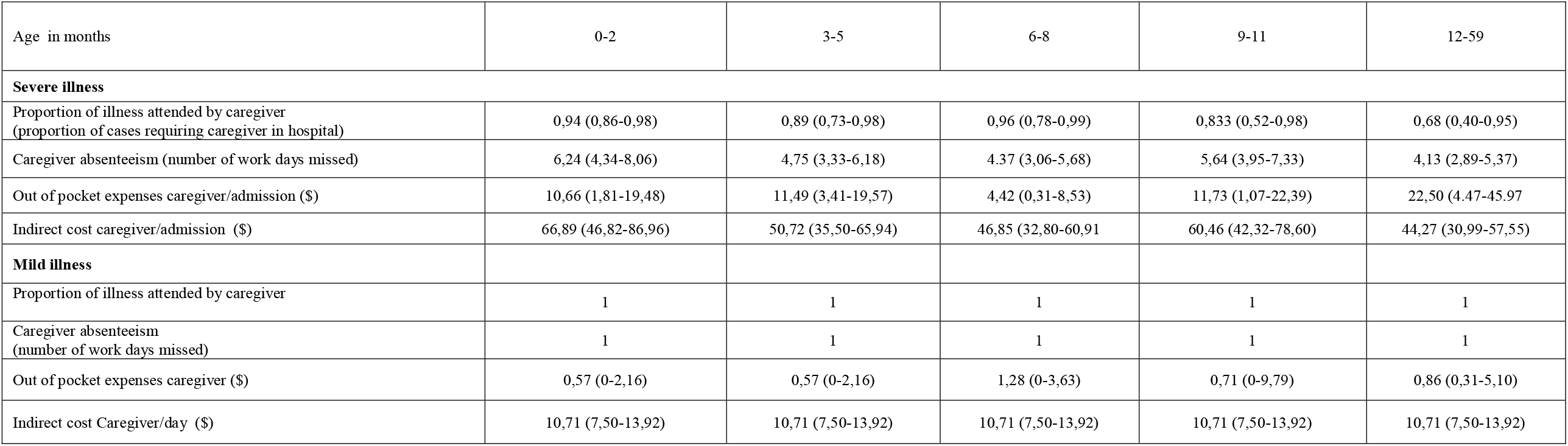
Mean caregiver (out of pocket and indirect) costs per episode for medically attended RSV-associated mild and severe illness in children aged <5 years, South Africa, 2013-2015

| Age in months | 0-2 | 3-5 | 6-8 | 9-11 | 12-59 |
| --- | --- | --- | --- | --- | --- |
| **Severe illness** |  |  |  |  |  |
| Proportion of illness attended by caregiver<br>(proportion of cases requiring caregiver in hospital) | 0,94 (0,86-0,98) | 0,89 (0,73-0,98) | 0,96 (0,78-0,99) | 0,833 (0,52-0,98) | 0,68 (0,40-0,95) |
| Caregiver absenteeism (number of work days missed) | 6,24 (4,34-8,06) | 4,75 (3,33-6,18) | 4,37 (3,06-5,68) | 5,64 (3,95-7,33) | 4,13 (2,89-5,37) |
| Out of pocket expenses caregiver/admission (\$) | 10,66 (1,81-19,48) | 11,49 (3,41-19,57) | 4,42 (0,31-8,53) | 11,73 (1,07-22,39) | 22,50 (4,47-45,97) |
| Indirect cost caregiver/admission (\$) | 66,89 (46,82-86,96) | 50,72 (35,50-65,94) | 46,85 (32,80-60,91) | 60,46 (42,32-78,60) | 44,27 (30,99-57,55) |
| **Mild illness** |  |  |  |  |  |
| Proportion of illness attended by caregiver | 1 | 1 | 1 | 1 | 1 |
| Caregiver absenteeism<br>(number of work days missed) | 1 | 1 | 1 | 1 | 1 |
| Out of pocket expenses caregiver (\$) | 0,57 (0-2,16) | 0,57 (0-2,16) | 1,28 (0-3,63) | 0,71 (0-9,79) | 0,86 (0,31-5,10) |
| Indirect cost Caregiver/day (\$) | 10,71 (7,50-13,92) | 10,71 (7,50-13,92) | 10,71 (7,50-13,92) | 10,71 (7,50-13,92) | 10,71 (7,50-13,92) |

### Cost burden

The estimated mean annual cost of RSV-associated Illness in children aged <5 years is $137 204 393 of which 76% ($111 742 713) are healthcare system incurred, 7% ($8 881 612) are out-of-pocket expenses and 17% ($24 225 801) are indirect costs. Thirty-three percent ($45 652 677) of the total costs were in the 0-2-month age group (Table 3).

**Table 3:** Mean annual national cost burden of RSV-associated mild and severe illness in children aged <5 years, South Africa, 2013 to 2015.

|  | Age group in months |  |  |  |  |  |
| --- | --- | --- | --- | --- | --- | --- |
| Costs in USD (\$) | 0-2<br>N (% of total) | 3-5<br>N (% of total) | 6-8<br>N (% of total) | 9-11<br>N (% of total) | 12-59<br>N (% of total) | Total costs<br>< 5 years |
| **Severe Illness** |  |  |  |  |  |  |
| **Total** | **41 082 860 (43)** | 17 668 519 (19) | 9 910 518 (10) | 5 458 436 (6) | 20 851 616 (22) | 94 971 949 |
| **Direct** | **38 801 193 (43)** | 16 584 414 (18) | 9 288 632 (10) | 5 150 035 (6) | 20 084 878 (22) | 89 909 152 |
| **Healthcare** | **38 435 544 (43)** | 16 357 168 (18) | 9 161 493 (10) | 5 092 867 (6) | 20 002 604 (22) | 89 049 676 |
| **Out of pocket** | **365 649 (43)** | 227 246 (26) | 127 139 (15) | 57 168 (7) | 82 274 (10) | 859 476 |
| **Indirect** | **2 281 667 (45)** | 1 084 104 (21) | 621 885 (12) | 308 401 (6) | 766 738 (15) | 5 062 795 |
| **Mild illness** |  |  |  |  |  |  |
| **Total** | 1 262 599 (5) | 830 645 (3) | 347 148(1) | 1 157 516 (4) | **23 918 961 (87)** | 27 516 869 |
| **Direct** | 859 569 (4) | 565 498(2) | 244 517 (1) | 980 019 (4) | **20 251 158 (88)** | 22 900 761 |
| **Healthcare** | 841 433(4) | 553 566 (2) | 239 899 (1) | 972 032 (4) | **20 086 107 (89)** | 22 693 037 |
| **Out of pocket** | 18 136 (9) | 11 932 (6) | 4 618 (2) | 7 987 (4) | **165 051 (79)** | 207 724 |
| **Indirect** | 403 030 (9) | 265 148 (6) | 102 631 (2) | 177 497 (4) | **3 667 802 (79)** | 4 616 108 |
| **Non-medically attended** |  |  |  |  |  |  |
| **Total** | 3 307 218 (21) | 1 546 779 (10) | 1 546 779 (10) | 601 610 (4) | **8 603 377 (55)** | 15 605 763 |
| **Direct** | 1 057 580 (13) | 510 314 (6) | 510 314 (6) | 291 669 (4) | **5 763 349 (71)** | 8 133 226 |
| **Indirect** | 2 249 638 (30) | 1 036 465 (14) | 1 036 465(14) | 309 941 (4) | **2 840 028 (38)** | 7 472 537 |
| **Total** |  |  |  |  |  |  |
| **Total** | 45 652 677 (33) | 20 045 943 (15) | 10 914 258 (8) | 7 217 562 (5) | 53 373 953 (39) | 137 204 393 |
| **Direct** | 40 718 342 (34) | 17 660 226 (15) | 9 724 649 (8) | 6 421 723 (5) | 46 099 385 (38) | 120 624 325 |
| **Healthcare** | 39 276 977 (35) | 16 910 734 (15) | 9 401 392 (8) | 6 064 899 (5) | 40 088 711 (36) | 111 742 713 |
| **Out of pocket** | 1 441 365 (16) | 749 492 (8) | 323 257 (4) | 356 824 (4) | 6 010 674 (68) | 8 881 612 |
| **Indirect** | 4 934 335 (30) | 2 385 717 (14) | 1 189 609 (7) | 795 839 (5) | 7 274 568 (44) | 28 225 801 |
| **Years of life lost** | 17812 | 10431 | 5185 | 2684 | 3513 | 39625 |

### Severe illness costs

The cost of RSV-associated severe illness was highest in the younger infants (0-2 months), with 43% ($38 435 544/$89 049 676) of the healthcare system costs in this age group. Similarly, the highest proportions of the total out-of-pocket and indirect costs were in this age group; 43% ($365 649 /$859 476) and 43% ($2 281 667 /$5 062 795), respectively. Healthcare system costs for severe illness were relatively low in the 12-59 age group with 22% ($20 002 604 /$89 049 676) of the healthcare costs in this age group (Table3).

### Mild illness costs

The largest cost burden of mild illness was in the 12-59-month age group, with 89% ($20 086 107 /$22 693 037) of the healthcare system costs in this age group. Similarly, the cost burden in this age group for out-of-pocket costs and indirect costs are highest in this age group ($165 051 and $3 667 802, respectively) (Table 3).

### Non-medically attended illness

Non-medically attended illness costs are higher in the older age group, similar to mild illness, with 55% ($8 603 377 /$15 605 763) of costs in the 1-4-year age group. Non-medically attended illness accounted for 11% ($15 605 763/$137 204 393) of the total costs of RSV-associated illness.

### Years of life lost

We estimated years of life lost for deaths for children < 5 years with RSV-associated illness to be 39625 years of which 45% (17812) occurred in the 0-2-month age group (Table 3).

## Discussion

In this analysis we were able to quantify the costs and cost-burden of RSV-associated illness, specifically the high costs and cost-burden of severe illness (hospitalized) in the youngest infants (aged 0-2 months), largely driven by high healthcare system costs in this age group. Costs are high throughout the first 6 months of life, the age groups targeted for new interventions. In contrast, for milder and non-medically attended illness, the majority of costs were in children aged 1-4 years, with significant costs of non-medically attended illness. This is the first analysis of the cost and cost burden of RSV-associated illness using pathogen specific cost data derived from South Africa and provides important data to support cost effectiveness models for interventions such as maternal vaccination and long-acting monoclonal antibodies aimed at young infants.

There are few data to compare to our analysis as few studies include finer age bands (most study provide costs for children aged <5 years) and most studies document the costs of pneumonia rather than pathogen-specific costs^13,14^. Two studies conducted in South Africa, one describes the cost per episode of influenza-associated severe illness (all ages) as $685,60 and the second study reports the cost per episode as $1139-for all cause pneumonia in children^12,14,15^. In this analysis we describe the average cost of RSV-associated severe illness for children aged <5 years of $681,42. In our study were describe a wide range of costs in the first five years of life and more specially in the first three months of life, allowing a more specific cost-effectiveness model for the age-specific interventions.

In a review article that included data from low and middle income countries (LMIC) compared with high income countries (HIC), the cost per episode of pneumonia ranged from $10-$1648 for inpatient care in LMIC (including data from South Africa), similar to our estimates of $591,44-$921,66 for RSV-associated inpatient care. Similar to our study the costs for pneumonia were driven by the duration of hospitalisation^13^. This review found a wide range of costs between different LMIC and differences in management of hospitalized patients vs community care for pneumonia between countries, supporting the need for country-specific cost estimates to drive country-level decision making.

Stenberg et al updated the WHO-CHOICE models in 2018 with data from 30 countries to develop a robust model to estimate country-level costs^16^. Our average daily cost for hospitalization in the <5 year age group ($136,81) was similar to estimates for Ecuador ($151,41) and Algeria ($131,41) and substantially more than estimates for Mozambique ($9,76). Although models like these are a valuable, specifically for countries that do not have the resources to conduct costing studies, the extrapolations used may not reflect the actual costs in that setting.

Healthcare is free to all children aged <5 years in South Africa, which means that the South Africa government budget carries most of the burden of the costs of RSV-associated illness. The South African healthcare budget in 2016, was 38 563 billion Rand ($33,53 billion)^17^. Although the cost of RSV-associated illness is <1% of the total healthcare budget, the cost saved by the introduction of a cost-effective intervention would release substantial funds to spend on strengthening other infant and child health prevention strategies^18^.

Although out of pocket and indirect cost are smaller proportions of the total costs of RSV-associated illness, the percentage of household income for these costs is high, specifically in a setting with high unemployment. In addition, the non-medically attended illness costs, specifically in the 1-4-year age group adds additional pressure on cash-strapped households.

Our study has several limitations. The costs of long-term sequelae such as repeated wheezing and loss of lung function following RSV infection are not captured in our study. In the South African Drakenstein cohort study a significant proportion of children experienced repeat admission and numerous follow up visits to clinics for these complications^19^. The incidence rate ratio for RSV-associated recurrent wheezing requiring hospitalization following RSV-LRTI was 1,48 (95% CI 1,30-1,68), suggesting a significant burden and cost-burden. We did not estimate the direct and indirect costs of RSV-associated deaths, for example additional loss of income and costs of a funeral. Our costing data collected in a single year may not reflect small changes in individual costs due to medical inflation or smaller fluctuations over the 3-year period.

significant

## Conclusion

The highest costs to the healthcare system for RSV-associated pediatric illness in South Africa were in very young infants resulting in the highest cost burden in this age group; therefore, use of these higher per illness cost estimates for interventions targeting young infants with either maternal vaccine or longer acting MAB may result in higher quality cost-effectiveness analyses. The substantial cost of RSV-associated mild illness and non-medically attended illness in older children aged 1-4 years suggests early childhood vaccination against RSV may also be a cost-effective intervention.

## Data Availability

All data produced in the present study are available upon reasonable request to the authors

